# Sharp Increases in Drug Overdose Deaths Among High-School-Age Adolescents During the US COVID-19 Epidemic and Illicit Fentanyl Crisis

**DOI:** 10.1101/2021.12.23.21268284

**Authors:** Joseph Friedman, Morgan Godvin, Chelsea Shover, Joseph P. Gone, Helena Hansen, David Schriger

## Abstract

Although overdose deaths in the US have increased exponentially for the past four decades, these shifts have historically affected adults, while pediatric overdose rates remained stable. However, this may be changing, given that the illicit drug supply has become increasingly hazardous in recent years, as illicitly-manufactured-fentanyls (IMFs) and other synthetic opioid and benzodiazepine analogues are increasingly sold as heroin and counterfeit prescription pills. We calculated drug overdose deaths per 100,000 population by 5-year age groups for the 2010-2021 period. For high-school-aged adolescents (age 14-18), we stratified rates by race/ethnicity, census region, associated substance, and ICD-10 cause-of-death intent categories. Adolescent overdose mortality saw a sharp increase between 2019 and 2020, from 2.35 per 100,000 to 4.58 per 100,000, representing a 94.3% increase, the largest percent increase of any 5-year age group. American Indian or Alaska Native (AIAN) adolescents, Latinx adolescents, and adolescents in the West census region were disproportionately affected, overdose death rates 2.15, 1.31, and 1.68 times the national average in 2021, respectively. Trends were driven by fatalities involving IMFs, which nearly tripled from 2019 to 2020, and represented 76.6% of adolescent overdose deaths in 2021. Sharp increases in adolescent drug overdose deaths, despite flat or declining drug use rates, and no increase in deaths from alcohol or most drugs, reinforce that rising fatalities are likely driven by an increasingly toxic, IMF-contaminated drug supply. Rising racial disparities in overdose require a prevention approach that ameliorates deep-seated social and economic inequalities as well as poor access to mental and physical healthcare and social services for AIAN and Latinx adolescents. Our results should also be understood in the context of rising rates of adolescent mental illness during the COVID-19 pandemic. These findings highlight the urgent need for accurate, harm-reduction-oriented education for early adolescents about the risks of an evolving drug supply, as well as greater access to naloxone and services that check drugs for the presence of IMFs.

## Introduction

The United States is in the midst of an unprecedented drug overdose crisis, recently crossing the milestone of 100,000 deaths in a 12-month period^1^. Although overdose deaths in the US have increased exponentially for the past four decades, these shifts have historically affected adults, while pediatric overdose rates remained stable^2^. The illicit drug supply has become increasingly hazardous in recent years, as illicitly-manufactured-fentanyls (IMFs) and other synthetic opioid and benzodiazepine analogues are increasingly sold as heroin and counterfeit prescription pills^3^. Although the number of adolescents using drugs reportedly declined in 2021^4^, media accounts of overdose deaths among high-school-age adolescents have become more frequent, often implicating counterfeit pills containing IMFs.

## Methods

We calculated drug overdose deaths per 100,000 population by 5-year age groups for the 2010-2021 period using data from the National Center for Health Statistics (see appendix). Records from 2020-2021 were provisional and may underestimate final mortality. Data from 2021 corresponded to January-May and were scaled proportionally to represent annual death counts and rates. For high-school-aged adolescents (age 14-18), we stratified rates by race/ethnicity, census region, associated substance, and ICD-10 cause-of-death intent categories. Analyses were conducted using R version 4.0.3. This work was deemed exempt from review by the University of California, Los Angeles, as it deals with anonymized and publicly available records.

## Results

Between 2010 and 2018, adolescents aged 14-18 had stable overdose mortality, substantially below the high and rising rates seen among the general population (supplemental Table). However, adolescents saw a sharp increase between 2019 and 2020, from 2.35 per 100,000 to 4.58 per 100,000, representing a 94.3% increase (Figure 1A). This was much larger than the percent increase observed among the all-age population in 2020, of 30.6% (Figure 1B). 14–18-year-old-individuals saw the highest percent increase in 2020 of any 5-year age group for whom trends could be assessed (Figure 1C). Overdose death rates among adolescents rose further in the first 5 months of 2021 to 5.50 per 100,000.

**Figure 1.**
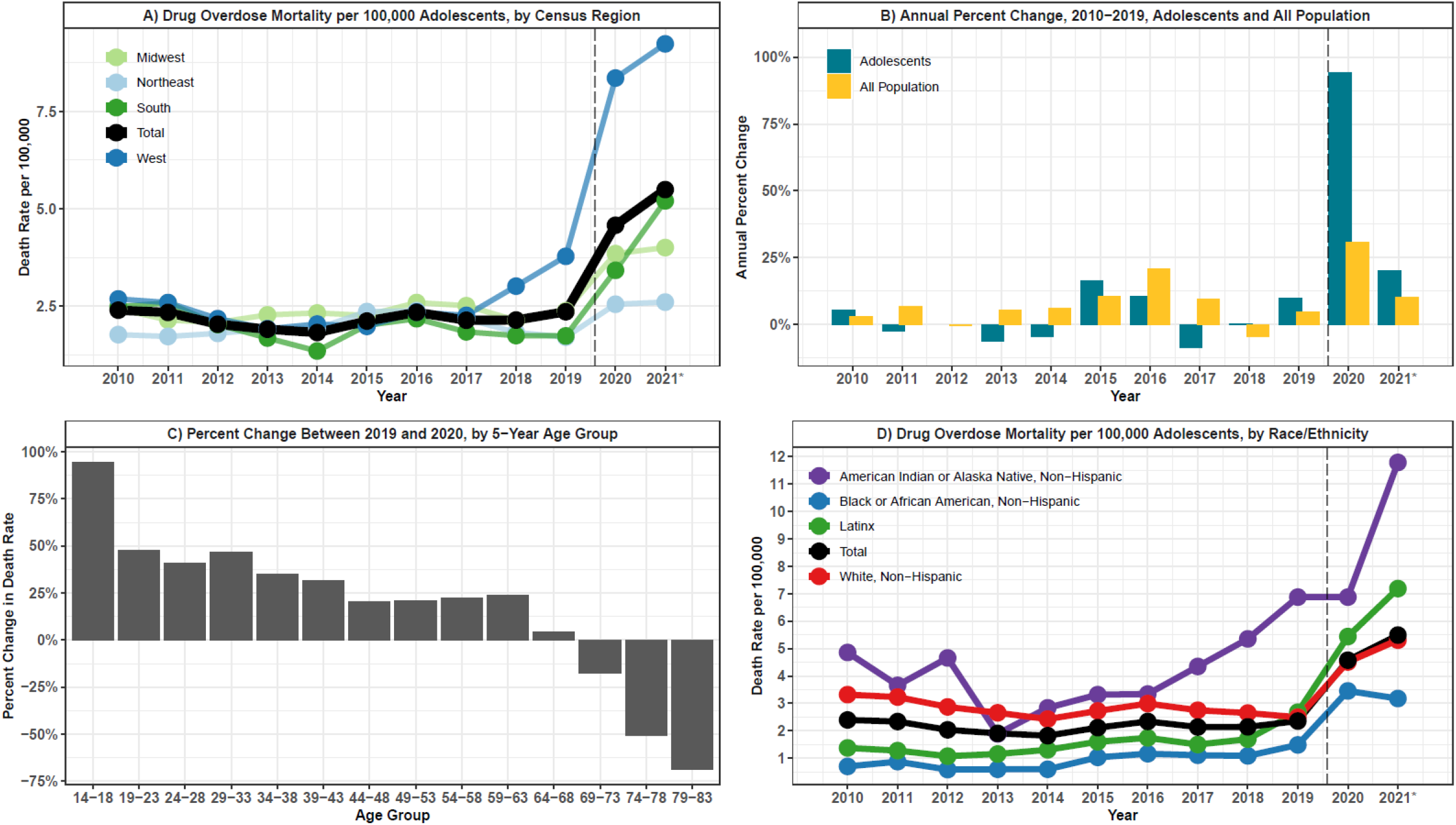
Adolescent Overdose Deaths, 2010-2021. Drug overdose deaths among high-school age adolescents (age 14-18) are shown per 100,000 individuals of the same age by race/ethnicity (A) and US Census Region (B). Year-to-year percent increases are shown for adolescents aged 14-18 and compared to the all-population figures (C). The percent change in drug overdose deaths between 2019 and 2020 is shown for 5-year age groups from ages 14-18 through ages 79-83. A vertical dashed line separates trends before and during the COVID-19 pandemic (A-C). *2021 refers to January-May 2021, and rates have been annualized.

Stratified by race/ethnicity, American Indian or Alaska Native (AIAN) adolescents saw disproportionate overdose mortality, especially after 2017. In provisional data from January-May 2021 overdose mortality in this group was 2.15-fold higher than the national average (Figure 1D). Latinx adolescents were also disproportionately affected, with a rate 1.31 times greater than the national average in 2021. Although adolescents in all census regions saw sharp increases in 2020-2021, those in the West were disproportionately affected, with an overdose death rate 1.68 times the national average. (Figure 1A).

Increases in adolescent overdose deaths can be attributed to rising fatalities involving IMFs and other synthetic opioids, which nearly tripled from 2019 to 2020, and represented 76.6% of all deaths by 2021 (Figure 2A). Throughout available data, most adolescent overdose deaths were categorized as unintentional, with a minority categorized as suicides, representing 84.5% and 13.4% of deaths in 2021, respectively (Figure 2B).

**Figure 2.**
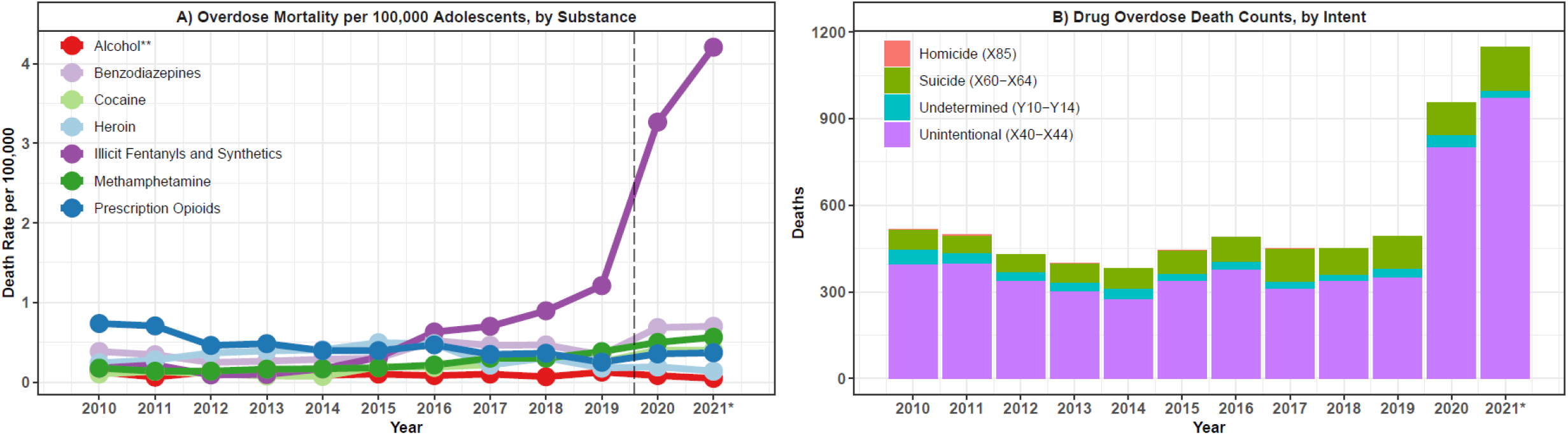
Substances-Involved and Intent of Adolescent Overdose Deaths, 2010-2021. Overdose deaths among high-school-aged adolescents (age 14-18) are shown per 100,000 individuals of the same age by substance used (A) and ICD-10 drug-related cause of death intent (B). A vertical dashed line separates trends before and during the COVID-19 pandemic (A). *2021 refers to January-May 2021, and rates and counts have been annualized. **Alcohol-related overdose deaths are shown here for comparative purposes but are not included in drug overdose rates shown elsewhere in the analysis, including Figure 1.

## Discussion

For decades, overdose mortality among high-school-aged adolescents remained stable and substantially lower than rates among the general population. However, beginning in 2020 they saw the largest annual percent change of any age group. The predominance of adolescent overdose deaths on the West coast may be related to the region’s high prevalence of counterfeit oxycodone and benzodiazepine pills containing high quantities of IMFs^3^. Sharp increases in adolescent drug overdose deaths, despite flat or declining drug use rates, and no increase in deaths from alcohol or most drugs, reinforce that rising fatalities are likely driven by an increasingly toxic, IMF-contaminated drug supply.

We noted sharp racial disparities for AIAN adolescents, of larger magnitude than those seen among adults in this population^5^. Disproportionately high trends among Latinx adolescents stand in contrast to relatively lower rates among Latinx adults^5^. Rising racial disparities in overdose require a prevention approach that ameliorates deep-seated social and economic inequalities as well as poor access to mental and physical healthcare and social services for AIAN and Latinx adolescents.

Our results should be understood in the context of rising rates of adolescent mental illness during the COVID-19 pandemic^6^. Although detected suicides remain a small proportion of adolescent overdose deaths, sharply increasing youth suicidal ideation may be a key contextual factor. These findings highlight the urgent need for accurate, harm-reduction-oriented education for early adolescents about the risks of an evolving drug supply, as well as greater access to naloxone and services that check drugs for the presence of IMFs.

## Data Availability

All data prodiced in the present study are publicly available online at: https://wonder.cdc.gov/mcd-icd10.html and https://wonder.cdc.gov/mcd-icd10-provisional.html. The authors may be contacted for assistance.

## Supplemental Methods

### Data Preparation and Definitions

1. Final drug overdose mortality and population counts were obtained for the 2010-2019 period from the CDC Wonder Final Multiple Cause of Death Platform (https://wonder.cdc.gov/mcd-icd10.html)
2. Provisional drug overdose counts for January 2020 – May 2021 were obtained from the CDC Wonder Provisional Multiple Cause of Death Platform (https://wonder.cdc.gov/mcd-icd10-provisional.html).
3. Drug overdose mortality rates per 100,000 individuals were calculated by 5-year age groups from ages 14-18 through ages 79-83. Individuals under age 14 were excluded due to insufficient overdose deaths to calculate a stable rate. Individuals over 83 were excluded due to missing population numbers in the Wonder platform.
4. Additionally, for individuals aged 14-18, overdose deaths per 100,000 individuals were calculated stratified by race/ethnicity, census region, substance-involved, and ICD-10 cause-of-death intent categories.
5. Drug overdoses were defined by the following underlying cause of death ICD-10 codes:

6. Age-specific populations served as the denominator and overdose death counts served as the numerator for the calculation of rates. Population data corresponding to the provisional period (2020-2021) were not available, so values from 2019 were used for these years, assuming no population change. Population change has been very small year-to-year, however this assumption could lead to slightly elevated rates.
7. ‘Latinx’ individuals were defined as any persons for which ethnicity was given as Hispanic or Latino, regardless of race. All other race/ethnicity groups were defined as individuals given to be of each race, who had their ethnicity listed as “non-Hispanic.”
8. With the exception of alcohol overdose deaths, substance-specific drug overdose deaths were defined by the following multiple cause of death ICD-10 codes, given that the underlying cause of death was assigned to one of the aforementioned underlying cause of death codes:

|  |
| --- |
| X40 (Accidental poisoning by and exposure to nonopioid analgesics, antipyretics and antirheumatics) |
| X41 (Accidental poisoning by and exposure to antiepileptic, sedative-hypnotic, antiparkinsonism and psychotropic drugs, not elsewhere classified) |
| X42 (Accidental poisoning by and exposure to narcotics and psychodysleptics [hallucinogens], not elsewhere classified) |
| X43 (Accidental poisoning by and exposure to other drugs acting on the autonomic nervous system) |
| X44 (Accidental poisoning by and exposure to other and unspecified drugs, medicaments and biological substances) |
| X60 (Intentional self-poisoning by and exposure to nonopioid analgesics, antipyretics and antirheumatics) |
| X61 (Intentional self-poisoning by and exposure to antiepileptic, sedative-hypnotic, antiparkinsonism and psychotropic drugs, not elsewhere classified) |
| X62 (Intentional self-poisoning by and exposure to narcotics and psychodysleptics [hallucinogens], not elsewhere classified) |
| X63 (Intentional self-poisoning by and exposure to other drugs acting on the autonomic nervous system) |
| X64 (Intentional self-poisoning by and exposure to other and unspecified drugs, medicaments and biological substances) |
| X85 (Assault by drugs, medicaments and biological substances) |
| Y10 (Poisoning by and exposure to nonopioid analgesics, antipyretics and antirheumatics, undetermined intent) |
| Y11 (Poisoning by and exposure to antiepileptic, sedative-hypnotic, antiparkinsonism and psychotropic drugs, not elsewhere classified, undetermined intent) |
| Y12 (Poisoning by and exposure to narcotics and psychodysleptics [hallucinogens], not elsewhere classified, undetermined intent) |
| Y13 (Poisoning by and exposure to other drugs acting on the autonomic nervous system, undetermined intent) |
| Y14 (Poisoning by and exposure to other and unspecified drugs, medicaments and biological substances, undetermined intent) |

9. Intent was defined by ICD-10 drug/alcohol induced cause of death codes.
10. Data were visualized using R version 4.0.3

| Substance | Definition | Notes |
| --- | --- | --- |
| Alcohol | Underlying cause of death in the "Alcohol poisonings (overdose) (X45, X65, Y15)" drug/alcohol induced cause set. | Not included in total drug overdose rates. Shown here for comparative purposes. |
| Benzodiazepines | MCD code "T42.4 (Benzodiazepines)" |  |
| Cocaine | MCD Code "T40.5 (Cocaine)" |  |
| Heroin | MCD Code "T40.1 (Heroin)" |  |
| Illicit Fentanyl and Synthetics | MCD Code "T40.4 (Other synthetic narcotics)" | This category is known to represent almost entirely illicitly-manufactured fentanyl and other synthetic opioids. A very small percentage of this category now includes prescription fentanyl. |
| Methamphetamine | MCD Code "T43.6 (Psychostimulants with abuse potential)" | This category is known to represent almost entirely illicit methamphetamine, and is traditionally referred to as such. See: https://jamanetwork.com/journals/jamapsychiatry/fullarticle/2774859 |
| Prescription Opioids | MCD Code "T40.2 (Other opioids)" | This category refers almost entirely to prescription opioids, such as oxycodone, not including methadone or prescription fentanyl. |

### Methodological Considerations

1. Records from 2020-2021 are provisional and may underestimate final mortality rates.
2. Data from 2021 represent only January-May and could overestimate or underestimate subsequently observed trends representing the entire year.
3. Race/ethnicity can be incorrectly assigned in some death investigations. This is a well-documented issue for American Indian and Alaska Native individuals in particular, and should inspire caution in the interpretation of results.
4. These findings are descriptive in nature and cannot indicate causally if increases in overdose mortality were directly linked to the COVID-19 pandemic or any other factor.
5. For drug overdoses, as with nearly all causes of death, mortality rates remain much higher for adults than for pediatric populations. The key finding of this work include that adolescents aged 14-18 now represent the fastest growing segment of the drug overdose crisis in relative, year-to-year percent increase space. This is particularly notable after many years of flat trends. However, these findings should not discount the importance of drug overdose among adults, whose overall rates still remain higher. Rather, as has been seen for numerous demographic groups (e.g., Black communities in urban areas) early exponential increases may precede subsequent large increases in level. Therefore, there may be an opportunity now for early intervention among a segment of the population with rapidly growing overdose mortality. Many of these considerations can be seen in the primary results when shown numerically, as below:

| Year | Adolescents (14-18) |  |  |  | All Population |  |  |  |
| --- | --- | --- | --- | --- | --- | --- | --- | --- |
|  | Deaths | Death Rate per 100,000 | Percent Change | Absolute Change | Deaths | Death Rate per 100,000 | Percent Change | Absolute Change |
| 2010 | 518 | 2.4 | 5.4 | 0.1 | 38329 | 12.4 | 2.9 | 0.4 |
| 2011 | 497 | 2.3 | -2.4 | -0.1 | 41340 | 13.3 | 6.9 | 0.9 |
| 2012 | 429 | 2.0 | -12.9 | -0.3 | 41502 | 13.2 | -0.4 | 0.0 |
| 2013 | 399 | 1.9 | -6.4 | -0.1 | 43982 | 13.9 | 5.2 | 0.7 |
| 2014 | 382 | 1.8 | -4.5 | -0.1 | 47055 | 14.8 | 6.1 | 0.8 |
| 2015 | 445 | 2.1 | 16.2 | 0.3 | 52404 | 16.3 | 10.5 | 1.5 |
| 2016 | 491 | 2.3 | 10.6 | 0.2 | 63632 | 19.7 | 20.8 | 3.4 |
| 2017 | 449 | 2.1 | -8.7 | -0.2 | 70237 | 21.6 | 9.5 | 1.9 |
| 2018 | 450 | 2.1 | 0.2 | 0.0 | 67367 | 20.6 | -4.5 | -1.0 |
| 2019 | 492 | 2.4 | 9.9 | 0.2 | 70630 | 21.5 | 4.5 | 0.9 |
| 2020 | 956 | 4.6 | 94.3 | 2.2 | 92232 | 28.1 | 30.6 | 6.6 |
| 2021 | 1147 | 5.5 | 20.0 | 0.9 | 101460 | 30.9 | 10.0 | 2.8 |

